# Type 2 Diabetes Mellitus and Risk of Colorectal Cancer: A Systematic Review and Meta-Analysis

**DOI:** 10.64898/2026.11.13.26367733

**Authors:** Shengyang Wang, Anna Porter, Nianjun Liu, Lin Ge

## Abstract

Epidemiologic studies suggest that type 2 diabetes mellitus (T2DM) is associated with an increased risk of colorectal cancer (CRC). However, prior meta-analyses have relied heavily on studies published more than a decade ago and have often combined distinct epidemiologic effect measures. We conducted an updated systematic review and meta-analysis of observational studies published from January 2012 through January 2026 evaluating the association between T2DM and incident CRC. PubMed, Embase, Web of Science, CENTRAL, and Scopus were searched, and eligible studies reporting associations between T2DM and CRC were included. Effect estimates were synthesized separately using random-effects models to calculate pooled estimates with 95% confidence intervals (CIs). Sex-specific and leave-one-out sensitivity analyses were also conducted. Fourteen studies were included, comprising nine cohort and five case-control studies; six reported odds ratios (ORs), seven reported hazard ratios (HRs), and one reported a standardized incidence ratio (SIR). T2DM was associated with higher odds of incident CRC (OR, 1.28; 95% CI, 1.13–1.46) and a higher hazard of incident CRC (HR, 1.27; 95% CI, 1.23–1.31). The study reporting an SIR also showed higher CRC incidence among individuals with T2DM (SIR, 1.40; 95% CI, 1.39–1.45). Sex-specific analyses generally showed positive associations in both females and males, and sensitivity analyses supported the robustness of the pooled findings. Overall, T2DM was associated with an increased risk of incident CRC, with a consistent direction of association across distinct epidemiologic effect measures. These findings provide updated evidence supporting consideration of T2DM as a marker of increased CRC risk and may inform CRC prevention strategies among individuals with T2DM.

## Introduction

Type 2 diabetes mellitus (T2DM) accounts for more than 90% of diabetes cases worldwide.^1,2^ In 2024, an estimated 589 million adults were living with diabetes, a number projected to reach 853 million by 2050.^2,3^ T2DM has been associated with an increased risk of several cancers, including liver, pancreatic, endometrial, and colorectal cancers.^4,5^ Colorectal cancer (CRC) is the third most commonly diagnosed cancer and the second leading cause of cancer death worldwide, accounting for approximately 1.9 million new cases and 900,000 deaths in 2022.^6^ CRC incidence has also increased among younger adults in recent decades. In the United States, an estimated 158,850 new CRC cases and 55,230 CRC deaths are projected in 2026.^7^ Given the growing burden of T2DM and CRC, clarifying the association between these conditions may inform cancer risk stratification, prevention, screening, and early detection.^6^

Several biological mechanisms may contribute to the association between T2DM and colorectal carcinogenesis.^8–10^ Insulin resistance and compensatory hyperinsulinemia may promote cellular proliferation and inhibit apoptosis through insulin and insulin-like growth factor signaling pathways.^11–14^ Chronic low-grade inflammation, oxidative stress, altered adipokine profiles, and metabolic dysregulation associated with T2DM may also contribute to tumor initiation and progression. ^15–19^ In addition, T2DM and CRC share several risk factors, including older age, obesity, physical inactivity, and unhealthy dietary patterns, complicating the estimation of an independent association.^20–23^

Although previous systematic reviews and meta-analyses have generally reported a positive association between T2DM and the risk of incident CRC,^24^ important limitations remain. Prior meta-analyses relied heavily on studies published more than a decade ago and did not incorporate recent evidence.^25,26^ Moreover, some previous meta-analyses combined odds ratios (ORs), hazard ratios (HRs), and standardized incidence ratios (SIRs) into a single pooled estimate.^26^ These measures represent different epidemiologic contrasts and often arise from different study designs; pooling them without distinction may therefore obscure clinically and methodologically relevant differences.^27^ Sex may also modify the association between T2DM and CRC, potentially through differences in hormonal, metabolic, behavioral, and health care–related factors.^28,29^ However, sex-specific associations have not been comprehensively characterized in published meta-analyses.

To address these gaps, we conducted an updated systematic review and meta-analysis of studies published from January 2012 through January 2026 to evaluate the association between T2DM and the risk of incident CRC. ORs, HRs, and SIRs were analyzed separately to preserve the interpretation of each effect measure. We also conducted sex-specific and sensitivity analyses to assess potential differences between males and females and to evaluate the robustness of the findings.

## Methods

### Search strategy

This meta-analysis was reported in accordance with the Preferred Reporting Items for Systematic Reviews and Meta-Analyses (PRISMA) guidelines.^30^ The literature search was conducted in January 2026, in PubMed, Embase, Web of Science, the Cochrane Central Register of Controlled Trials (CENTRAL), and Scopus. Studies published between January 2012 and January 2026 were considered for inclusion. The January 2012 cutoff was selected based on the evidence window of a previous meta-analysis that included studies published through 2012.^25,26^ Search terms, including controlled vocabulary and free-text keywords, were combined using Boolean operators. The detailed search strategy is provided in Supplementary Table S1. Only studies published in English were included. Reference lists of included studies were also manually searched to identify additional relevant articles.

### Inclusion and exclusion criteria

Eligibility criteria were defined using the PECO framework: Population (P), individuals at risk of developing CRC; Exposure (E), diagnosed T2DM; Comparator (C), individuals without T2DM or a general-population reference group; and Outcome (O), incident CRC. Studies were included in the meta-analysis if they met all the following criteria: (1) published between January 2012 and January 2026; (2) evaluated the association between T2DM and the risk of incident CRC; (3) reported effect estimates, including ORs, HRs, or other relative risks, with corresponding 95% CIs; and (4) provided multivariable-adjusted estimates. Studies were excluded if they met any of the following criteria: (1) conference abstracts, reviews, or summary reports; (2) included only individuals with type 1 diabetes; or (3) reported outcomes other than CRC incidence, such as survival or mortality.

### Data extraction and quality assessment

Two investigators (S.W. and L.G.) independently screened the literature according to the predefined inclusion and exclusion criteria. For each included study, the following information was extracted: first author, publication year, study design, geographic region, source population, study period, sample size, covariates adjusted for, and reported effect estimates with corresponding CIs. When multiple articles analyzed the same or overlapping study samples, only the study with the largest sample size was included to avoid duplication of data.

Study quality was assessed independently by two investigators (S.W. and L.G.) using the 9-star Newcastle-Ottawa Scale (NOS).^31^ The NOS evaluates studies across three domains: selection, comparability, and outcome (for cohort studies) or exposure (for case-control studies). Studies with scores ≥ 7 out of 9 were considered high quality.^31^ Quality assessment results were summarized separately for cohort and case-control studies in Supplementary Tables S2 and S3, respectively.

### Statistical analysis

The association between T2DM and the risk of incident CRC was evaluated using the effect estimates reported in the included studies, together with corresponding 95% confidence interval (CIs). Because ORs, HRs, and SIRs represent distinct epidemiologic measures, they were analyzed separately. Pooled effect estimates and 95% CIs were calculated using a random-effects model, with the between-study variance estimated using the DerSimonian-Laird method. Between-study heterogeneity was assessed using Cochran’s Q test, the I² statistic, and the between-study variance (τ²).

When a study did not report an overall effect estimate but provided sex-specific estimates, the sex-specific estimates were combined using a fixed-effect model to derive a study-level overall estimate. A p-value < 0.05 for Cochran’s Q test was considered indicative of statistically significant heterogeneity. I² values of < 25%, 25–50%, 50–75%, and > 75% were interpreted as indicating low, moderate, substantial, and considerable heterogeneity, respectively. Sex-specific meta-analyses were conducted using the same random-effects modeling approach for studies reporting estimates separately for males and females.

### Sensitivity analysis and publication bias

Sensitivity analyses were performed using a leave-one-out approach, in which each study was sequentially excluded and the pooled effect estimate with 95% CIs was recalculated. Publication bias was assessed to evaluate potential small-study effects and selective reporting. Funnel plots were used to examine potential asymmetry. Egger’s regression test was also performed to evaluate publication bias, with p < 0.05 indicating statistically significant asymmetry. All analyses were performed using the *metafor*package in R (version 4.5.3).

## Results

### Study selection and characteristics

The study selection process is shown in Figure 1.

**Figure 1.**
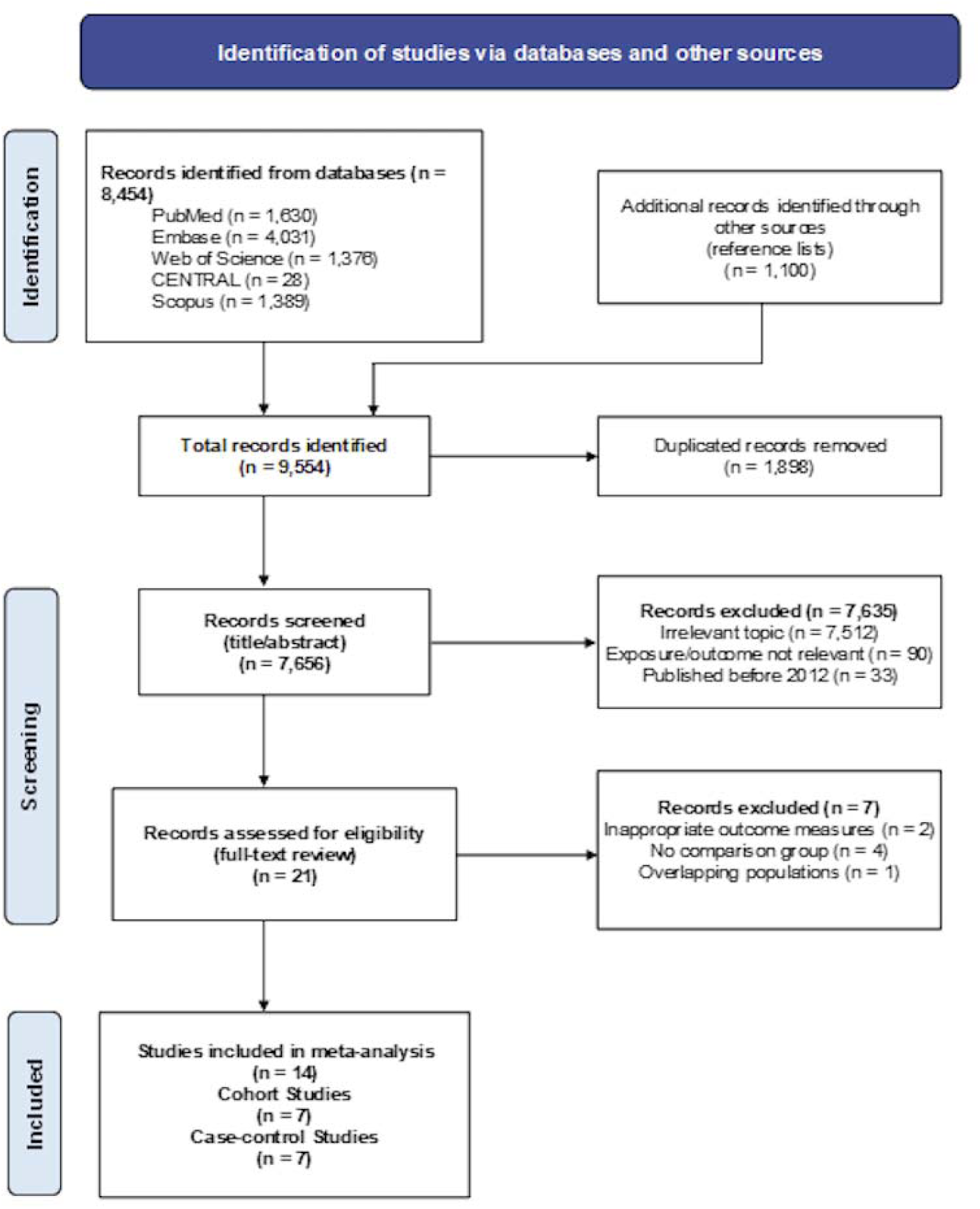
PRISMA flow diagram of the literature search and study selection process.

A total of 9,554 records were identified through searches of PubMed, Embase, Web of Science, the Cochrane Central Register of Controlled Trials (CENTRAL), and Scopus. After 1,898 duplicate records were removed, 7,656 records remained for title and abstract screening. Of these, 7,635 were excluded because they were not relevant to the research question (n = 7,512), did not evaluate the exposure or outcome of interest (n = 90), or did not meet the prespecified publication-date criterion (n = 33). Twenty-one articles underwent full-text review, of which 14 met the eligibility criteria and were included in the meta-analysis.

Among the 14 included studies, six reported odds ratios (ORs), seven reported hazard ratios (HRs), and one reported a standardized incidence ratios (SIRs). The characteristics of the included studies are summarized in Table 1.

**Table 1.** Characteristics of studies included in the meta-analysis.

| **First author** | **Year** | **Title** | **Study design** | **Region** | **Source population** | **Study period** | **Total participants** | **Confounders** | **Adjusted effect estimate (95% CI)** |
|---|---|---|---|---|---|---|---|---|---|
| Li | 2022 | Type 2 Diabetes and Risk of Early-Onset Colorectal Cancer. | Case-control | United States | U.S. commercially insured adults (IBM MarketScan database) | 2006–2015 | 6001 cases, 52,104 controls | Age, sex, insurance duration, region, drug coverage, employment, residence, insurance type, Charlson comorbidity index, IBD, obesity, family history of GI cancer, colonoscopy, FOBT. | OR = 1.24 (95% CI: 1.09–1.41) |
| Rezazadeh | 2024 | Effect of diabetes mellitus type 2 and sulfonylurea on colorectal cancer development: a case-control study. | Case-control | Iran | Hospital-based population (patients ≥45 years, Tehran, Iran) | 2019–2023 | 684 cases and 810 controls | Age, sex, aspirin, metformin, insulin, smoking, sulfonylurea | OR = 0.67 (95% CI: 0.37–1.20) |
| Wankhede | 2025 | Diabetes and Colorectal Cancer Risk and Survival According to Tumor Immunity Status. | Case-control | Germany | Population-based (DACHS study, adults ≥30 years, Germany) | 2003–2021 | 2,362 cases and 2,362 controls | Age, sex, education, alcohol, smoking (pack-years), BMI, red meat intake, physical activity, aspirin, statins, antihyperglycemic use, endoscopy, family history of CRC, medical examination history | OR = 1.39 (95% CI: 1.17–1.66) |
| Cavicchia | 2013 | Racial disparities in colorectal cancer incidence by type 2 diabetes mellitus status. | Cohort | United States | Medicaid beneficiaries aged ≥30 years with ≥12 months Medicaid eligibility | 1990–2007 | 91,836 participants (6,006 with T2DM; 85,681 without T2DM) | Age, sex, race, marital status, hypertension, Medicaid eligibility duration | OR = 1.20 (95% CI: 0.90–1.58) |
| Rosato | 2016 | Type 2 Diabetes, Antidiabetic Medications, and Colorectal Cancer Risk: Two Case-Control Studies from Italy and Spain. | Case-control | Italy and Spain | Italy: hospital-based controls; Spain: population-based controls | 2007–2013 | 1,147 cases and 1,594 controls | Age, sex, education, smoking, alcohol, BMI, physical activity, statins, aspirin | OR = 1.21 (95% CI: 0.95–1.55) |
| Woo | 2016 | Diabetes Mellitus and Site-specific Colorectal Cancer Risk in Korea: A Case-control Study | Case-control | Korea | Hospital-based (cases) and health screening controls | 2010–2013 | 1,070 cases and 2,775 controls | Age, BMI, alcohol, diet (overcooked meat), physical activity | OR = 1.62 (95% CI: 1.28–2.06)<sup>*</sup> |
| Ma | 2018 | Type 2 diabetes and risk of colorectal cancer in two large U.S. prospective cohorts. | Cohort | United States | Nurses’ Health Study (women) and Health Professionals Follow-up Study (men) | 1980–2012 (women); 1986–2012 (men) | 134,763 (87,523 women; 47,240 men) | Age, race, family history of diabetes, aspirin use, BMI, family history of CRC, endoscopy/sigmoidoscopy, smoking, alcohol, physical activity, total calorie intake, folate, calcium, vitamin D, processed meat, red meat, (plus hormone use in women) | HR = 1.25 (95% CI: 1.09–1.44)<sup>*</sup> |
| Overbeek | 2019 | Sex- and site-specific differences in colorectal cancer risk among people with type 2 diabetes. | Cohort | Netherlands | PHARMO GP Database linked with Netherlands Cancer Registry | 2006–2014 | ~270,000 (≈55,000 T2D + ≈215,000 controls) | Age, socioeconomic status (SES), aspirin, NSAIDs, statins, antihypertensives, hormone replacement therapy | HR = 1.29 (95% CI: 1.13–1.47)<sup>*</sup> |
| Mäkinen | 2025 | The association of type 2 diabetes and its complications with the risk of colorectal cancer. | Cohort | Finland | Finnish population-based cohort (Cancer Registry + Care Register) | 2000–2017 | 2,543,600 (171,789 with T2D) | Age, calendar period, sex, overweight/obesity, alcohol-related disorders | HR = 1.26 (95% CI: 1.20–1.32) |
| de Kort | 2016 | Diabetes mellitus type 2 and subsite-specific colorectal cancer risk in men and women: results from the Netherlands Cohort Study on diet and cancer. | Cohort | Netherlands | Netherlands Cohort Study on diet and cancer | 1986–2003 | Total participants = 120,852 | Age, BMI, waist proxy (clothing size), physical activity, smoking, alcohol, diet, family history of CRC | HR = 1.11 (95% CI: 0.82–1.48)<sup>*</sup> |
| Peeters | 2015 | The risk of colorectal cancer in patients with type 2 diabetes: associations with treatment stage and obesity. | Cohort | United Kingdom | Clinical Practice Research Datalink (CPRD) | 1987–2012 | Diabetic patients: n = 300,039; reference cohort: n = 300,039 | Age, sex, smoking, alcohol, statin use (± sensitivity with obesity) | HR = 1.26 (95% CI: 1.18–1.33) |
| Oh | 2020 | The Combined Impact of Chronic Kidney Disease and Diabetes on the Risk of Colorectal Cancer Depends on Sex: A Nationwide Population-Based Study | Cohort | South Korea | National Health Insurance Service–Health Examination Cohort | 2002–2013 | CKD−/DM+ disease group (n = 17,700) | Age, sex, income, BMI, smoking, alcohol, exercise, CCI score, family history, medication | HR = 1.34 (95% CI: 1.16–1.55) |
| Lawler | 2023 | Type 2 Diabetes and Colorectal Cancer Risk. | Cohort | United States | Southern Community Cohort Study | 2002–2018 | 25□992 participants with diabetes, 28□605 participants without diabetes | Age, sex, race, SES, screening, smoking, alcohol, BMI, family history | HR = 1.47 (95% CI: 1.21–1.79) |
| Chen | 2025 | Family history of diabetes mellitus and risk of colorectal cancer | Cohort | Sweden | Swedish nationwide registers | 1964–2018 | 11,784,567 individuals (668,348 with DM) | Age, sex, calendar period, SES, residence, obesity, alcoholism, COPD | SIR = 1.40 (95% CI: 1.39–1.45) |

Abbreviations: OR, odds ratio; HR, hazard ratio; SIR, standardized incidence ratio; CI, confidence interval.
<sup>\*</sup> Effect estimate calculated by the authors based on data reported in the original study.

Overall, 13 of 14 included studies reported effect estimates greater than 1.0, suggesting a positive association between T2DM and incident CRC. Among the six studies reporting ORs, five reported estimates consistent with higher odds of incident CRC, whereas Rezazadeh et al., 2024 reported a nonsignificant inverse association (OR = 0.67, 95% CI: 0.37–1.21). All seven studies reporting HRs yielded estimates greater than 1.0, indicating a higher hazard of incident CRC among individuals with T2DM. Similarly, the single study reporting an SIR found a higher incidence of CRC in individuals with T2DM than in the reference population (SIR = 1.40, 95% CI: 1.39–1.45). Study quality was evaluated using the NOS, with detailed assessments presented in Supplementary Tables S2 and S3. All included studies were classified as high quality based on their NOS scores.

### Pooled estimates of the association between T2DM and the risk of incident CRC

The pooled analysis of odds ratios showed that T2DM was associated with higher odds of incident CRC (OR = 1.28, 95% CI: 1.13–1.46, *p* < 0.001). Moderate between-study heterogeneity was observed (I² = 49.3%), although Cochran’s Q test was not statistically significant (p > 0.05). Similarly, the pooled hazard ratio indicated that T2DM was associated with a higher risk of incident CRC (HR = 1.27, 95% CI: 1.23–1.31, *p* < 0.001), with no evidence of between-study heterogeneity (I² = 0.0%, *p* > 0.05) (Figure 2).

**Figure 2.**
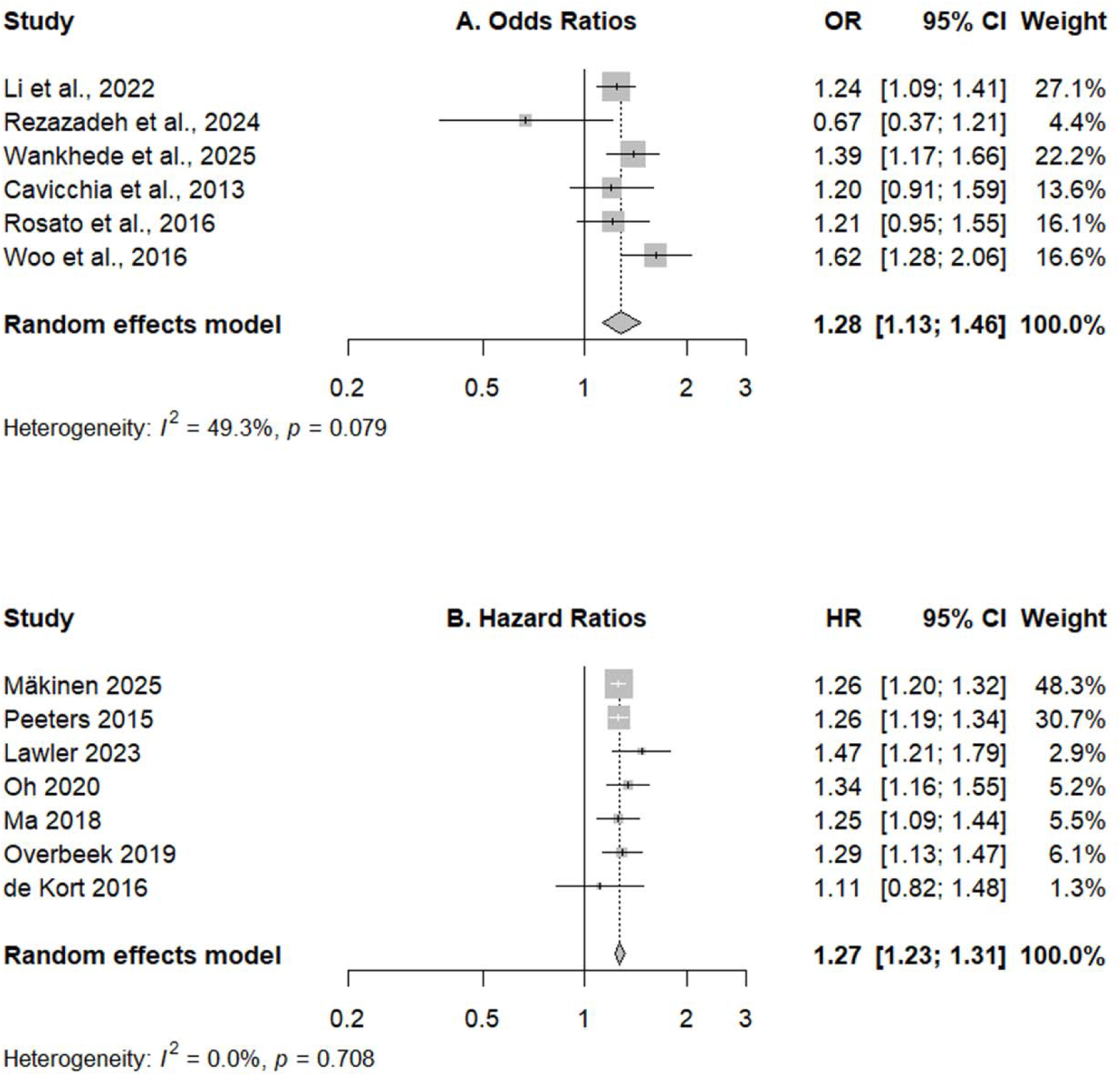
Forest plots of the association between type 2 diabetes mellitus and colorectal cancer risk. (A) Pooled odds ratios (ORs) and 95% confidence intervals (CIs). (B) Pooled hazard ratios (HRs) and 95% confidence intervals (CIs).

### Sex-specific analyses

Sex-specific analyses were conducted for studies reporting ORs and HRs. Two studies reporting ORs were included in the sex-specific analyses. Among females, the pooled OR was 1.48 (95% CI: 0.98–2.23, *p* > 0.05), with substantial between-study heterogeneity (I² = 62.9%), although Cochran’s Q test was not statistically significant (*p* > 0.05). Among males, T2DM was associated with higher odds of incident CRC (OR = 1.34; 95% CI: 1.09–1.64, *p* = 0.006), with moderate heterogeneity (I² = 42.1%, *p* > 0.05) (Figure 3A and 3B).

**Figure 3.**
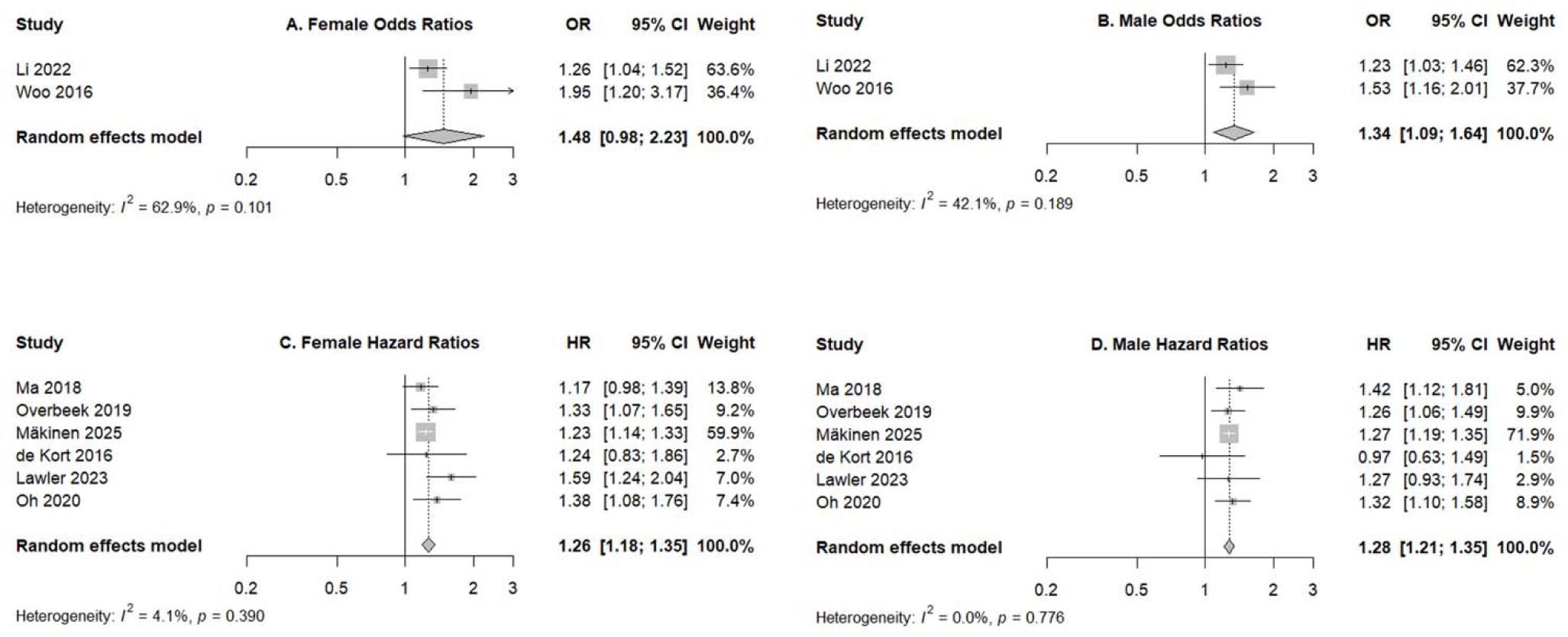
Forest plots of sex-stratified analyses of the association between type 2 diabetes mellitus and the risk of incident colorectal cancer. (A) Female odds ratios (ORs) and 95% confidence intervals (CIs). (B) Male odds ratios (ORs) and 95% confidence intervals (CIs). (C) Female hazard ratios (HRs) and 95% confidence intervals (CIs). (D) Male hazard ratios (HRs) and 95% confidence intervals (CIs).

Six studies reporting HRs were included in the sex-specific analyses. T2DM was associated with a higher risk of incident CRC in both females (HR = 1.26; 95% CI: 1.18–1.35, *p* < 0.001) and males (HR = 1.28; 95% CI: 1.21–1.35, *p* < 0.001). Between-study heterogeneity was low among females (I² = 4.1%, *p* > 0.05) and was not observed among males (I² = 0.0%, *p* > 0.05) (Figure 3C and 3D). One study reported sex-specific SIRs and found a higher incidence of CRC among both males (SIR = 1.50, 95% CI: 1.41–1.49) and females (SIR = 1.40, 95% CI: 1.34–1.42).^32^

### Sensitivity analyses and publication bias

Leave-one-out sensitivity analyses were conducted for the OR and HR meta-analyses (Supplementary Figure S1). In the OR analyses, the pooled estimates ranged from 1.24 to 1.32 after the sequential exclusion of individual studies, and all remained statistically significant. The heterogeneity (I² values) ranged from 19.7% to 58.2%, with the lowest heterogeneity observed after exclusion of Rezazadeh et al. (2024). In the HR analyses, pooled estimates ranged from 1.26 to 1.28 after the sequential exclusion of individual studies, which remained statistically significant and showed no evidence of heterogeneity (I²=0.0% across all iterations). Sensitivity analyses of the sex-specific HR estimates likewise yielded stable findings for both males and females. Sensitivity analyses were not performed for the sex-specific OR estimates because only two studies were available.

Publication bias was assessed using funnel plots and Egger’s regression test (Supplementary Figure S2). Egger’s test provided no significant evidence of funnel plots asymmetry in either the OR or HR meta-analysis (all p-values > 0.05), and visual inspection of the funnel plots revealed no marked asymmetry. Similar findings were observed in the sex-specific HR analyses. Nevertheless, because relatively few studies were included in each analysis, assessments of small-study effects and publication bias should be considered exploratory and interpreted with caution.

## Discussion

In this updated meta-analysis of studies published from January 2012 through January 2026, we evaluated the association between T2DM and incident CRC by analyzing epidemiological effect measures separately and examining sex-specific associations. T2DM was consistently associated with higher odds and a higher risk of incident CRC. The single study reporting a standardized incidence ratio (SIR) also found a higher incidence of CRC among individuals with T2DM.^32^ Sex-specific estimates were generally positive in both women and men; the pooled HRs excluded the null in both groups, whereas the pooled OR excluded the null only among men.

Our findings are consistent in direction with those of previous meta-analyses but extend the evidence in both recency and analytic approach. By including studies published from January 2012 through January 2026, the present study provides a more current assessment of the association and enhances the relevance of the findings to contemporary epidemiologic and clinical contexts. A previous meta-analysis combined effect estimates from cohort and case-control studies, including ORs and HRs, as a common summary RR.^26^ Although these measures may provide broadly comparable estimates under certain conditions, they are not numerically interchangeable and retain distinct epidemiologic interpretations. We therefore pooled ORs and HRs separately and reported the SIR independently, thereby preserving the interpretation of each effect measure. The concordant direction of the measure-specific findings indicates that the positive association between T2DM and incident CRC remains evident in contemporary epidemiologic evidence.

Between-study heterogeneity differed between the OR and HR analyses. Moderate heterogeneity was observed among studies reporting ORs, whereas no between-study heterogeneity was observed among studies reporting HRs. This contrast may partly reflect differences in study design and methodological characteristics.^33^ Five of 6 studies contributing to the pooled OR used a case-control design,^34–38^ which may be more sensitive to variation in control selection, exposure ascertainment, and selection or recall bias. In contrast, all 7studies contributing to the pooled HR were large population-based cohort studies with longitudinal follow-up.^39–45^ Their larger sample sizes may also have contributed to the greater precision of the HR estimates. However, because relatively few studies were included in each analysis, the heterogeneity estimates themselves may be imprecise; an I² value of 0% should therefore not be interpreted as definitive evidence of complete homogeneity.

The leave-one-out sensitivity analyses further supported the stability of the pooled findings. Sequential exclusion of individual studies did not substantially alter the pooled estimates, and all pooled OR and HR estimates remained statistically significant. In the OR analysis, heterogeneity was lowest after the exclusion of Rezazadeh et al. (2024),^35^ which reported a nonsignificant inverse association. This hospital-based case-control study included 684 cases and 810 hospitalized controls, and its estimate changed from a positive unadjusted association (OR, 1.39; 95% CI, 1.07-1.81) to an inverse adjusted association (OR, 0.67; 95% CI, 0.37-1.20) after adjustment for demographic characteristics, smoking, aspirin use, and diabetes medications.^35^ Its divergence from the other studies may partly reflect its relatively small sample and use of hospitalized controls, which may have reduced statistical precision and increased susceptibility to selection bias. Because the adjusted 95% CI included the null, the inverse estimate should not be interpreted as evidence of a protective association. Neither visual inspection of the funnel plots nor Egger regression tests indicated funnel-plot asymmetry. However, fewer than 10 studies were included in each analysis, limiting the power to detect asymmetry; publication bias therefore cannot be excluded. All included studies were assessed as high quality according to the NOS, supporting the reliability of the pooled findings. Nevertheless, quality scores cannot exclude residual confounding, exposure misclassification, selection bias, or other sources of bias inherent in observational research.

Sex-specific analyses indicated positive association in both females and males. The pooled HRs showed statistically significant associations in both groups, with similar magnitudes of association. The pooled OR estimate among males was statistically significant, whereas the pooled OR among females was elevated but had a 95% confidence interval that included the null value. The latter finding may reflect the small number of contributing studies, substantial between-study heterogeneity, and limited statistical precision rather than the absence of an association. The single study reporting sex-specific SIRs found an increased incidence of CRC among both females and males.^32^ Although the HR and SIR estimates were higher among males, the pooled OR was numerically higher among females. Thus, the available evidence does not demonstrate a consistent sex-related pattern or provide sufficient support for effect modification by sex. Formal comparisons between sex-specific estimates were limited by the small number of studies and the absence of sufficiently detailed stratified data.

Several biological and behavioral factors could potentially contribute to sex-related variation in the association between T2DM and CRC.^46,47^ These include differences in sex hormones, body-fat distribution, insulin sensitivity, inflammatory responses, health behaviors, and participation in CRC screening.^46–48^ However, the present findings do not establish that any of these factors explain the observed differences. Additional sex-stratified epidemiological studies and mechanistic investigations are needed to determine whether the association between T2DM and CRC meaningfully differs between female and male.

Several additional limitations should be acknowledged. The number of studies available for some pooled analyses was relatively limited, particularly for OR and sex-specific analyses. Although leave-one-out sensitivity analyses suggested stable pooled estimates, the limited number of available studies should be considered when interpreting these findings. Sex-specific analyses were also constrained by the availability of reported data, which limited further evaluation according to geographic region, tumor location, and other study characteristics. In addition, only English-language publications were included, raising the possibility of language bias. Future studies with larger sample sizes and more detailed stratified reporting are needed to clarify the association between T2DM and the risk of incident CRC across populations and disease characteristics.

## Conclusion

This meta-analysis found that T2DM was associated with higher odds and hazard of incident CRC, with the SIR estimate showing the same direction of association. Sex-specific estimates were generally positive among both females and males, although the available evidence was insufficient to determine whether the association differed by sex. These findings provide updated evidence supporting consideration of T2DM as a marker of the increased risk of incident CRC and may help inform CRC prevention strategies among individuals with T2DM.

## Acknowledgements

We thank Thea Patricia Atwood for her assistance with database searches and literature review.

## Data Availability

The data underlying this article are available in the article and in its online supplementary material.

## Declaration of generative AI and AI-assisted technologies in the manuscript preparation process

During the preparation of this work the author(s) used Chatgpt in order to check grammars. After using this tool/service, the author(s) reviewed and edited the content as needed and take(s) full responsibility for the content of the published article.

