## Supplementary Material for "Type 2 Diabetes Mellitus and Risk of Colorectal Cancer: A Systematic Review and Meta-Analysis"

**Supplementary Materials**

**Table S1. Table S1. Detailed search strategies for PubMed, Embase, Web of Science, CENTRAL, and Scopus.**

| Databases | Search Strategy |
| --- | --- |
| PubMed | ("type 2 diabet*" OR T2DM OR "type II diabet*")  AND  ("colorectal cancer*" OR CRC) |
| Embase | ('type 2 diabetes mellitus'/exp  OR 'type 2 diabetes':ti,ab,kw  OR 'type ii diabetes':ti,ab,kw  OR t2dm:ti,ab,kw)  AND  ('colorectal cancer'/exp  OR 'colon cancer':ti,ab,kw  OR 'rectal cancer':ti,ab,kw) |
| Web of Science | ("type 2 diabetes" OR "type II diabetes" OR "type 2 diabetic") AND ("colorectal cancer" OR "colorectal neoplasm*" OR "colon cancer" OR "rectal cancer") |
| CENTRAL | ("type 2 diabetes" OR "type II diabetes" OR T2DM)  AND  ("colorectal cancer" OR "colorectal neoplasm*" OR CRC) |
| Scopus | TITLE-ABS-KEY("type 2 diabetes" OR "type II diabetes" OR T2DM)  AND  TITLE-ABS-KEY("colorectal cancer" OR "colon cancer" OR "rectal cancer") |

**Table S2. Newcastle–Ottawa Scale quality assessment of cohort studies included in the meta-analysis**

| Study | Selection (0 - 4) | Comparability (0 - 2) | Outcome (0 - 3) | Total Score (0 - 9) |
| --- | --- | --- | --- | --- |
| Cavicchia et al., 2013 | 4 | 2 | 2 | 8 |
| Ma et al., 2018 | 3 | 2 | 2 | 7 |
| Overbeek et al., 2019 | 4 | 2 | 2 | 8 |
| Mäkinen et al., 2025 | 4 | 2 | 3 | 9 |
| Peeters et al., 2015 | 4 | 2 | 3 | 9 |
| Lawler et al., 2023 | 4 | 2 | 3 | 9 |
| Chen et al., 2025 | 4 | 2 | 3 | 9 |

Abbreviations: NOS, Newcastle–Ottawa Scale.
Studies scoring 7–9 points were classified as good quality, 5–6 points as fair quality, and 0–4 points as poor quality.

**Table S3. Newcastle–Ottawa Scale quality assessment of case-control studies included in the meta-analysis.**

| Study | Selection (0 - 4) | Comparability (0 - 2) | Outcome (0 - 3) | Total Score (0 - 9) |
| --- | --- | --- | --- | --- |
| Li et al., 2022 | 4 | 2 | 2 | 8 |
| Rezazadeh et al., 2024 | 3 | 2 | 2 | 7 |
| Wankhede et al., 2025 | 4 | 2 | 3 | 9 |
| Rosato et al., 2016 | 4 | 2 | 3 | 9 |
| Woo et al., 2016 | 3 | 2 | 2 | 7 |

Abbreviations: NOS, Newcastle–Ottawa Scale.
Studies scoring 7–9 points were classified as good quality, 5–6 points as fair quality, and 0–4 points as poor quality.

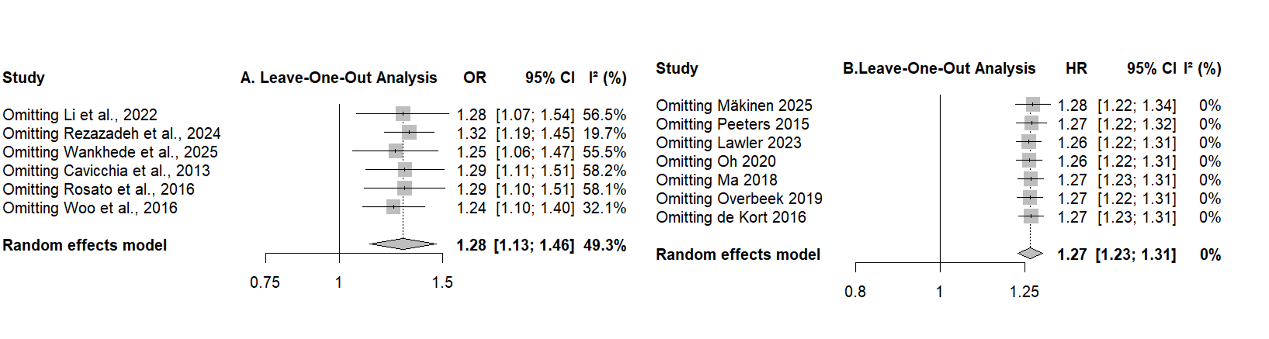

**Figure S1. Leave-one-out sensitivity analyses of the association between type 2 diabetes mellitus and colorectal cancer risk.**

(A) Odds ratio (OR) analysis. (B) Hazard ratio (HR) analysis.

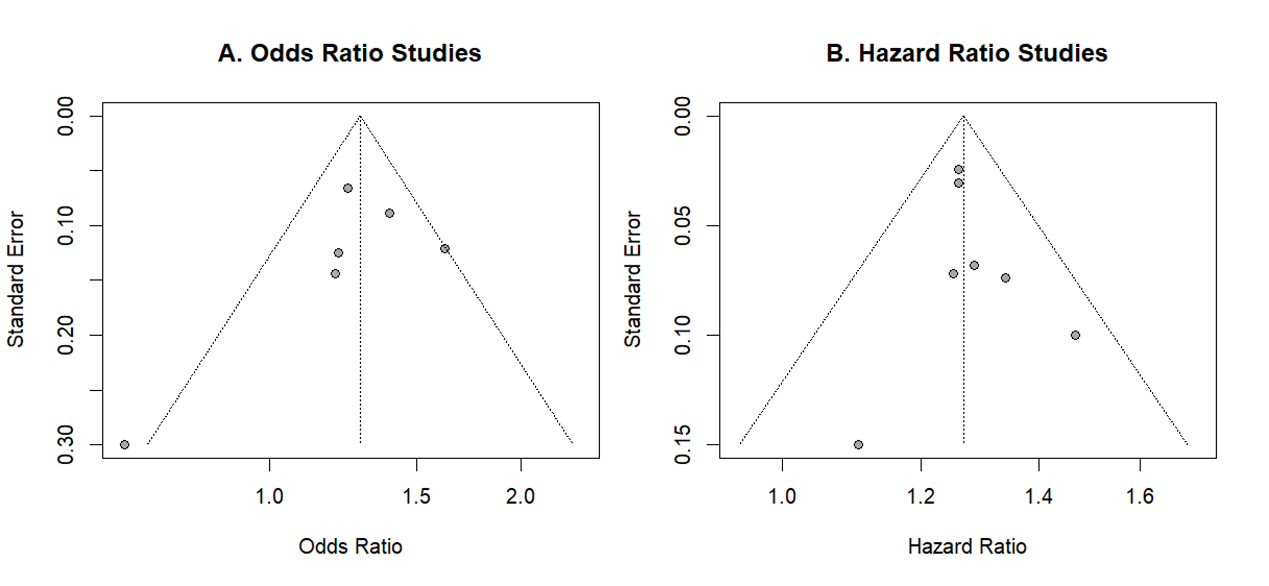

**Figure S2. Funnel plots for assessment of publication bias in studies evaluating the association between type 2 diabetes mellitus and colorectal cancer risk.**

(A) Studies reporting odds ratios (ORs). (B) Studies reporting hazard ratios (HRs).
